# Traumatic life events as predictors for depression in middle-aged men and women: A Finnish twin study

**DOI:** 10.1101/2024.04.18.24306034

**Authors:** Piirtola Maarit, Haravuori Henna, Kiviruusu Olli, Viertiö Satu, Suvisaari Jaana, Marttunen Mauri, Kaprio Jaakko, Korhonen Tellervo

**Affiliations:** Institute for Molecular Medicine Finland (FIMM), University of Helsinki, Helsinki, Finland; UKK Institute for Health Promotion Research, Tampere, Finland; Mental Health Team, Department of Public Health and Welfare, Finnish Institute for Health and Welfare (THL), Helsinki, Finland; Psychiatry, University of Helsinki and Helsinki University Hospital, Helsinki, Finland; Wellbeing services county of Lapland, Rovaniemi, Finland

## Abstract

**Background:** We examined the exposure to adulthood traumatic life events (TLEs) and analysed their associations with depression in women and men. Then we examined whether the associations of TLEs are independent of exposure loading and vulnerability including familial confounding.

**Methods:** Total of 8410 individuals (45% men, mean age 60 years) participated in the fourth survey of the population-based Finnish Twin Cohort conducted in 2011. Depression was assessed using the Center for Epidemiologic Studies Depression (CES-D) scale (cut-off value ≥20). Participants reported exposure to TLEs during adulthood. Logistic regression adjusted for multiple covariates was used as the individual-based analyses. The effect of shared familial factors was tested using conditional logistic regression in 399 twin pairs discordant for depression.

**Results:** Depression was more common in women (15%) than in men (11%). Men reported more traffic accidents (men: 11.8%, women: 7.4%), other serious accidents (11.8%, 5.8%), and violent crime (3.1%, 2.0%) whereas women reported more sexual assault (0.7%, 10.6%). Violent crime (Odds Ratio 3.86; 95% Confidence Intervals 2.59, 5.73), sexual assault (3.49; 2.67, 4.55), physical assault (3.10; 2.45, 3.93), and other serious accidents (1.36; 1.01, 1.85) were associated with depression. These associations, except other serious accidents, remained statistically significant after adjusting for multiple covariates including TLEs load and shared familial factors. The associations did not differ by sex.

**Conclusions:** Women and men differ in exposure to TLEs but, if exposed, they are equally vulnerable for depression. Because traumatic life events are robustly associated with depression, they should be considered in prevention and treatment.

## INTRODUCTION

Among adults, depression disorders are highly prevalent globally, and twice as prevalent in women than in men (Bebbington, 1996, Bromet *et al*., 2011, Salk, Hyde and Abramson, 2017, Steel *et al*., 2014), for reasons still poorly understood (Hyde, Mezulis and Abramson, 2008, Salk *et al*., 2017).

Depression is a complex multifactorial disorder (Bromet *et al*., 2011, Hyde *et al*., 2008, Kendler, Kuhn and Prescott, 2004a, Köhler *et al*., 2018, Markkula *et al*., 2015). It is, however, particularly common among individuals exposed to stressful and traumatic life events (TLEs) (Kessler, 1997, McCutcheon *et al*., 2009, Vibhakar, Allen, Gee and Meiser-Stedman, 2019, Xiang and Wang, 2021). Even though men report higher overall number of TLEs than women (Hatch and Dohrenwend, 2007) it is possible that women underreport some TLEs, such as domestic violence (Attila *et al*., 2023).

Overall, there are not many population-based studies analysing association of TLEs with depression separately in men and women (Kendler *et al*., 2004a, McLaughlin, Conron, Koenen and Gilman, 2010, Xiang and Wang, 2021). Even fewer studies have considered multiple potential confounders, such as personality and shared familial factors (Kendler and Gardner, 2014, Kendler, Thornton and Prescott, 2001). Despite many shared risk factors for depression and TLEs (Hatch and Dohrenwend, 2007, Otten *et al*., 2021), gender-based differences exist. It has been suggested that neuroticism and failures in interpersonal relationships play a larger role in the development of major depression in women whereas substance abuse, prior history of depression, and childhood sexual abuse play a greater role in men (Kendler and Gardner, 2014). Gender-based sensitivity for depression is, however, most likely modified by both socio-cultural factors as gender-based expectations and marital status, and traits like temperament and personality (Maji, 2018, Parker and Brotchie, 2010). Though, higher education, being married and having fewer previous TLEs have been found to protect against subsequent TLEs in general (Benjet *et al*., 2016), the evidence for TLEs as gender-based risks for depression is sparse. Generally, the relationship between TLEs and depression has been analysed in women only (Kendler, Kuhn and Prescott, 2004b) or gender has been adjusted for in the analyses (Richardson, Carr, Netuveli and Sacker, 2020).

There are several theories related to impact of TLEs on depression among men and women. According to exposure hypothesis (Denton, Prus and Walters, 2004), women and men differ by their exposure to TLEs which would then partly explain gender differences in depression. However, adverse childhood experiences (ACE) have been reported to associate equally with adult TLEs in both genders (Bürgin *et al*., 2021). Notably, even if exposure would differ, the same TLE might not be equally traumatic to everyone (Pearlin, Schieman, Fazio and Meersman, 2005), and not all individuals exposed to TLEs suffer from depression (Baldwin, Reuben, Newbury and Danese, 2019, McLaughlin *et al*., 2010). Therefore, gender, personality, socioeconomic factors, social relationships, exposure to ACE and genetic liability, including biological sex, are likely to confound or modify the complex aetiology of exposure to TLEs and of depression onset (Kendler *et al*., 2004a, Kessler, 1997, Luo, Zhang and Roberts, 2021, Pearlin *et al*., 2005). Hence, differences in depression have also been explained through interaction with individual-based vulnerability, such as sensitivity to adverse experiences (Bilodeau, Marchand and Demers, 2020, Piccinelli and Wilkinson, 2000). In addition, an exposure load (sensitivity/sensitization) hypothesis has been proposed (Hammen, Henry and Daley, 2000, McLaughlin *et al*., 2010) suggesting that individuals facing TLEs during adulthood are more sensitive for depression if they have also experienced ACEs (Bandoli *et al*., 2017, Kendler *et al*., 2004b, McLaughlin *et al*., 2010). Further, it has been reported that individuals who experience adversities, such as traumas, are at an increased risk of experiencing further adversities during their life span (Pearlin *et al*., 2005). Notably, also a dose-response relationship between stressors and depression has been shown indicating that more frequent or more severe stressors either in childhood or in adulthood lower the threshold for depression (Bandoli *et al*., 2017, Kendler *et al*., 2004b, McLaughlin *et al*., 2010).

Familial liability for depression cannot been ruled out in the development of depression after adverse life events (Zimmermann *et al*., 2008). Twin and family studies indicate a moderate heritability of depression (Flint and Kendler, 2014, Kendall *et al*., 2021, Polderman *et al*., 2015), which may be higher in women (Flint and Kendler, 2014). Likewise, there is evidence for a heritable component in trauma exposure (Lyons *et al*., 1993, Power *et al*., 2013), and this may be shared with the heritable component of depression (Coleman 2020). Therefore, in addition to individual-based risk factors for depression, familial factors including both childhood environmental factors and genetic vulnerability are potential confounders when analysing associations between TLEs and depression. For this purpose, twin studies provide a powerful design allowing control of familial factors (McGue, Osler and Christensen, 2010).

## AIMS

First, we report sex-differences in prevalence of depression based on the Center for Epidemiologic Studies Depression (CES-D) scale and in the exposure to TLE during adulthood. Second, we analyse sex differences in the associations of TLEs with depression. Third, we test if the associations are independent of variables increasing vulnerability for depression such as personality, socioeconomic factors, other negative life events, and ACEs, as well as familial factors including genetic liability.

## METHODS

### Participants

The fourth survey of the population-based Finnish Twin Cohort of like-sexed twin pairs was conducted in 2011 (Kaprio, 2013). Participation rate was 72% and the full data included 8,410 participants (46% men, age range: 53 to 68 years) (Figure 1a). We had information only on biological sex from the registered birth certificates. Gender was not inquired in the questionnaires of the twin cohort. Participant characteristics by sex and depressive symptoms are described in Table 1.

**Figure 1a.**
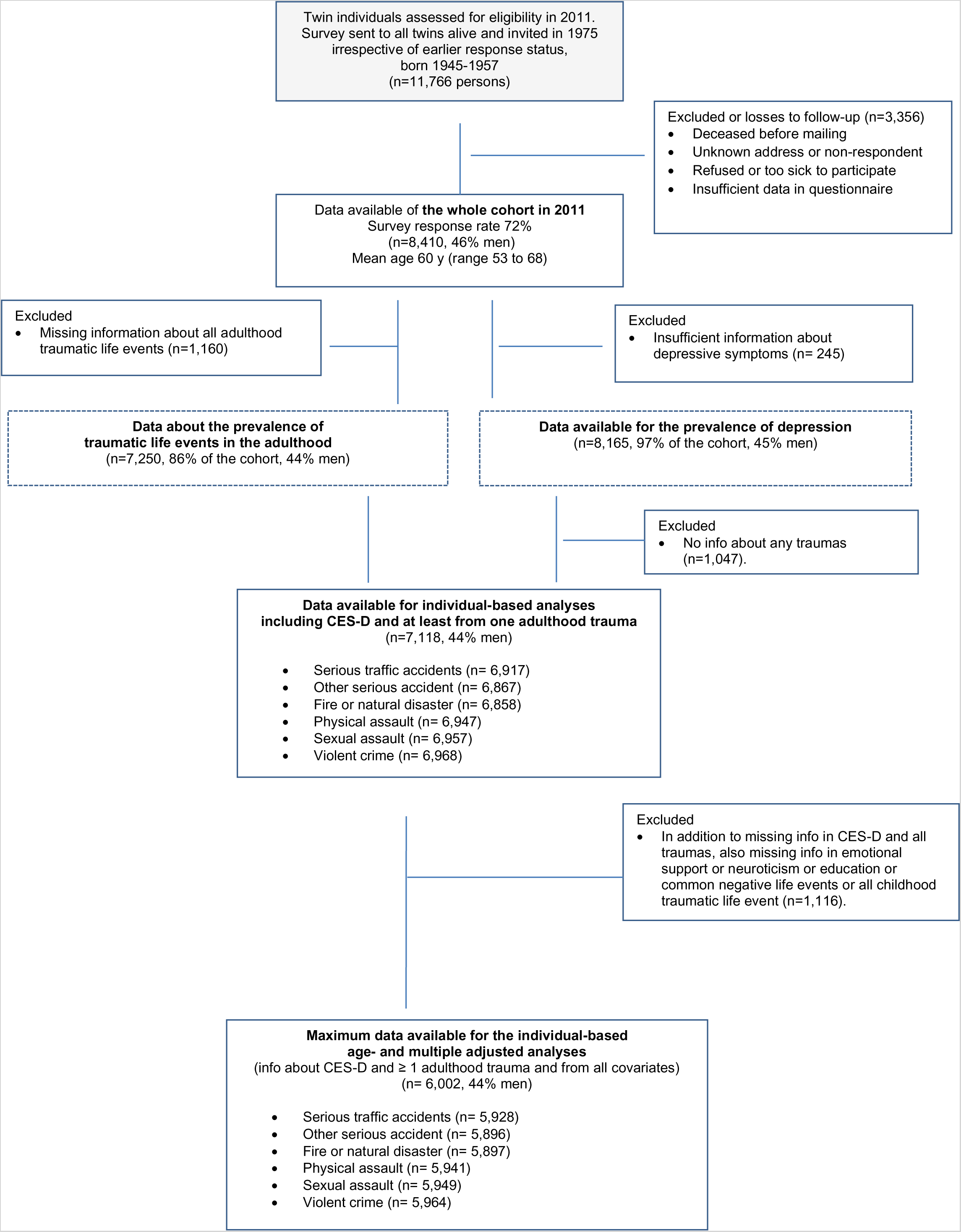
Flow chart of samples included in the individual-based analyses.

**Table 1.** Characteristics of participants (n=8,165) with CES-D information by sex in the Finnish Twin Cohort in 2011.

| **Depression status and background information from 2011** (if not mentioned otherwise) | **Men (n=3,634)** |  | **Women (n=4,531)** |  |
| --- | --- | --- | --- | --- |
|  | CES-D <20 | CES-D ≥20 | CES-D <20 | CES-D ≥20 |
| n (%) | 3,227 (88.8) | 407 (11.2) | 3,850 (85.0) | 681 (15.0) |
| **Emotional support, n (%)** |  |  |  |  |
| Strong support | 2,741 (84.9) | 273 (67.1) | 3,615 (93.9) | 526 (77.2) |
| Some support | 280 (8.7) | 65 (16.0) | 162 (4.2) | 79 (11.6) |
| Not any support | 185 (5.7) | 65 (16.0) | 56 (1.5) | 71 (10.4) |
| Missing info | 21 (0.7) | 4 (1.0) | 17 (0.4) | 5 (0.7) |
| **Marital status, n (%)** |  |  |  |  |
| Single | 324 (10.0) | 94 (23.1) | 412 (10.7) | 102 (15.0) |
| Married or living together | 2,550 (79.0) | 214 (52.6) | 2,636 (68.5) | 382 (56.1) |
| Divorced or widowed | 334 (10.4) | 94 (23.1) | 784 (20.4) | 195 (28.6) |
| Missing info | 19 (0.6) | 5 (1.2) | 18 (0.5) | 2 (0.3) |
| **Working status, n (%)** |  |  |  |  |
| At work | 1,653 (51.2) | 129 (31.7) | 2,131 (55.4) | 265 (38.9) |
| Retired | 793 (24.6) | 63 (15.5) | 849 (22.1) | 132 (19.4) |
| On disability pension | 403 (12.5) | 126 (31.0) | 231 (9.4) | 146 (21.4) |
| Out of work for other reasons | 372 (11.5) | 86 (21.1) | 496 (12.9) | 137 (20.1) |
| Missing info | 6 (0.2) | 3 (0.7) | 13 (0.3) | 1 (0.2) |
| **Exposure to serious adverse childhood experiences<sup>a</sup>, n (%)** |  |  |  |  |
| Not any | 2,329 (72.2) | 235 (57.7) | 2,905 (75.5) | 370 (54.3) |
| At least one | 533 (16.5) | 88 (21.6) | 531 (13.8) | 172 (25.3) |
| Missing info | 365 (11.3) | 84 (20.6) | 414 (10.8) | 139 (20.4) |
| **Age, mean (sd)** | 60.5 (3.7) | 60.2 (3.8) | 60.1 (3.8) | 59.9 (3.7) |
| **Education years until 1981, mean (sd)** | 8.7 (3.1) | 8.0 (2.6) | 9.0 (3.1) | 8.3 (2.8) |
| Missing info, n (%) | 12 (0.4) | 0 (0) | 4 (0.1) | 2 (0.2) |
| **Neuroticism sum score, mean (sd)** | 2.7 (2.2) | 6.1 (2.2) | 2.9 (2.1) | 6.3 (2.1) |
| Missing info, n (%) | 74 (2.3) | 13 (3.2) | 71 (1.8) | 17 (2.5) |
| **Sum of time-weighted negative life events, mean (sd)** | 6.1 (4.1) | 9.4 (5.1) | 6.6 (4.1) | 10.2 (5.2) |
| Missing info, n (%) | 94 (2.9) | 43 (10.6) | 123 (3.2) | 46 (6.8) |

CES-D=The Center for Epidemiological Studies-Depression.
<sup>a</sup> Traumatic life events in childhood: exposure to serious traffic accident, other serious accident, fire and natural disaster, physical assault, sexual assault and/or violent crime.

The prevalence of depression was calculated for 8,165 participants who completed the CES-D assessment (Figure 1a). The prevalence of TLEs was calculated for those 7,250 who had also reported exposure (yes/no) to at least one TLEs during adulthood (44% men). The Individual-based association analyses were performed for those 6,002 (44% men) participants who had information on depressive symptoms, TLEs and relevant covariates.

Within-pair data came from 2,253 same-sexed twin pairs (40% male pairs) where both twins had information about their depression status and at least one mention about their exposure to TLEs during adulthood (Figure 1b). We identified 399 pairs (35% male) who were discordant for depression (as defined below), of whom 264 were dizygotic (DZ) and 135 monozygotic (MZ).

**Figure 1b.**
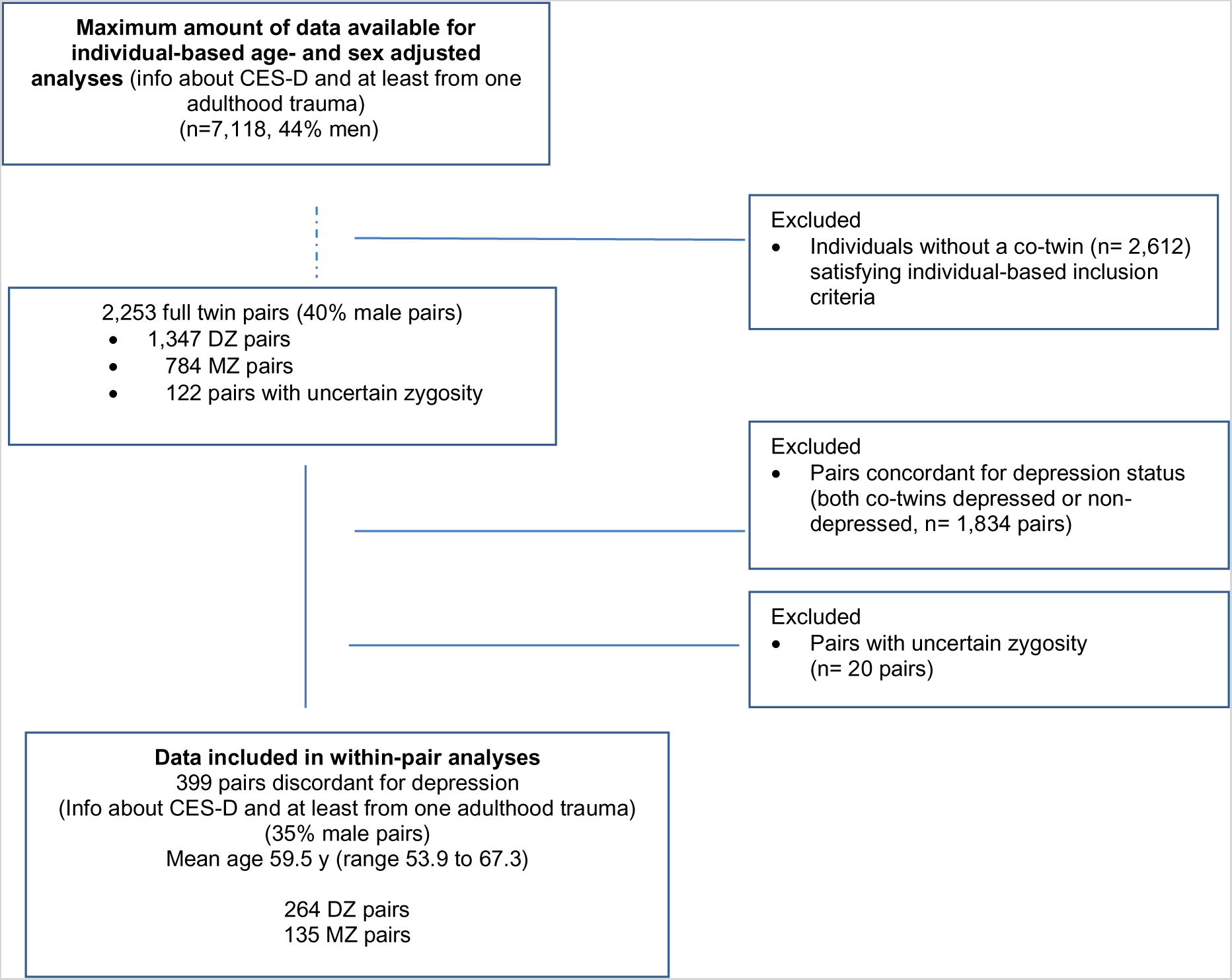
Flow chart of twin pairs included in the within-pair analyses.

### Ethics

All procedures contributing to this work comply with the ethical standards of the relevant national and institutional committees on human experimentation and with the Helsinki Declaration of 1975, as revised in 2008. The study protocol was approved by the Ethical Committee of the University of Helsinki, Department of Public Health. Analyses of the present study did not involve contact with the study participants. A returned and completed questionnaire was considered as consent from the participant. The study participants were provided with information about the study aims and their right to withdraw from the study at any time.

### Measures

#### Depression

Depressive symptoms were assessed using the sum score of the self-reported 20-item Center for Epidemiologic Studies Depression (CES-D) scale (range 0-60 points). A higher score reflects more current depressive symptoms (Radloff, 1977). If one or two items were missing, they were imputed by replacing the mean of the non-missing items before computing the sum score (616 participants, 7% of the total sample). The process is described in detail elsewhere (Piirtola *et al*., 2021). As the sum score was skewed with a floor effect (Fig S1), we coded depression as a dichotomous variable (yes/no) with a cut off value ≥20. This classifies an individual with a clinically relevant level of depression (Vilagut, Forero, Barbaglia and Alonso, 2016).

#### Traumatic life events

Exposure (yes/no) to traumatic life events (TLEs) during adulthood was assessed using the Trauma History Screen measure (Carlson *et al*., 2011). We asked exposure to six items: involvement in a serious traffic accident, involvement in any other serious accident, being a witness to a fire or natural disaster, being injured by a physical assault, being a victim of a sexual assault, and being a victim of or witness to a violent crime. Cumulative number of TLEs was calculated both as a sum and categorized as none, one or two, and at least three TLEs in adulthood. The corresponding items were asked from childhood and calculated both as cumulative sum and categorized ACE (none vs. at least one).

A total of 6,527 participants (90%) of those reporting information at least one TLE (n=7,268) had no missing information in any of their adulthood TLEs. In the whole cohort, missing information varied from 15.8% to 18.1% per TLE, with no sex-difference (Table 2). Persons with depression, irrespective of sex, had about twice as much missing information in each TLE than those without depression.

**Table 2.**
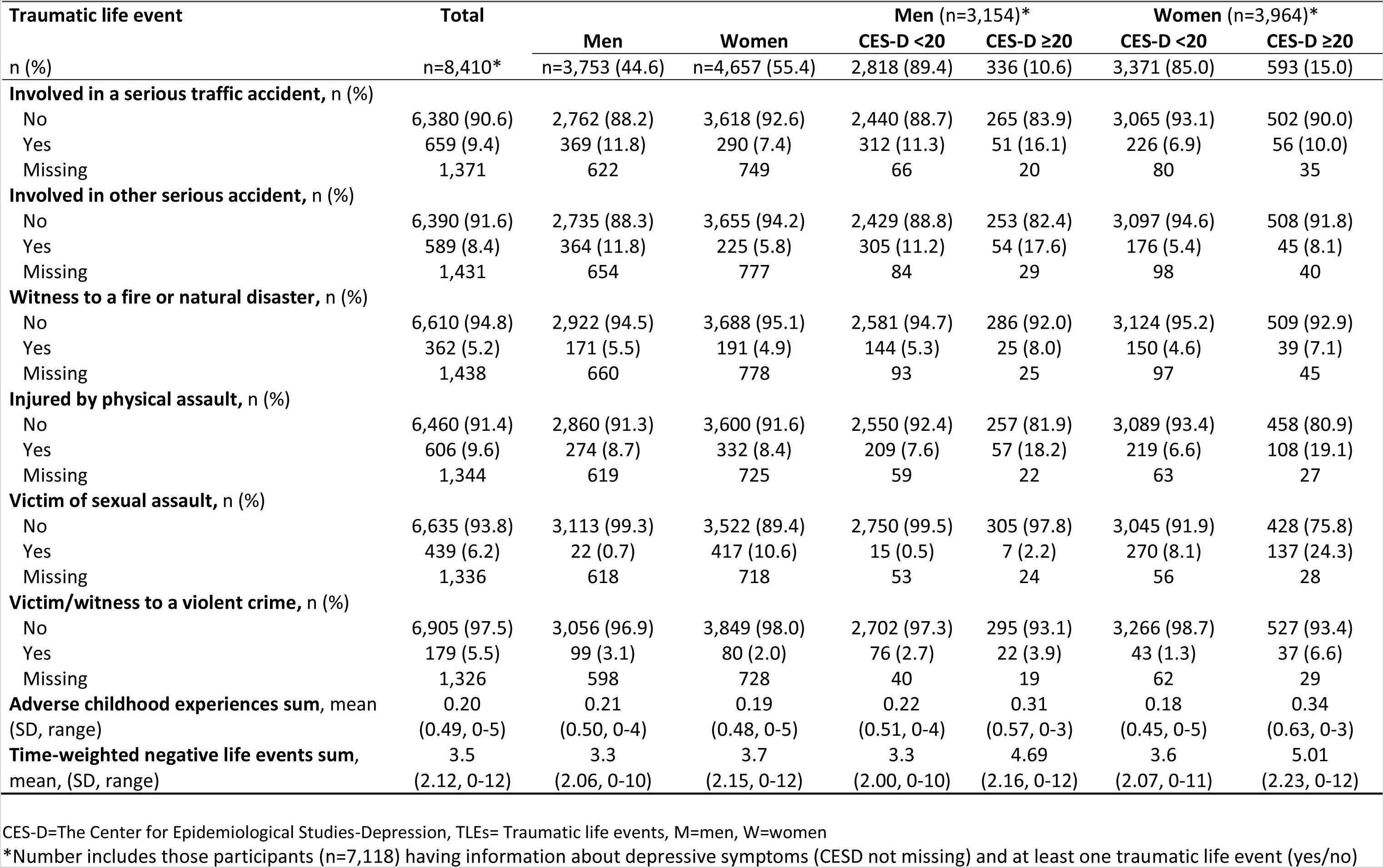
Prevalence of self-reported adulthood traumatic life events, adverse childhood experiences and negative life events in the whole cohort by sex (n=8,410) and by sex and depressive symptoms (n=7,118) * in the Finnish Twin cohort in 2011.

#### Background variables and covariates

Register data provided sex and date of birth. Age was computed using date of response and birthdate. Relevant covariates associated with depression and TLEs were chosen based on previous literature (Bromet *et al*., 2011, Kendler *et al*., 2004a, Kendler *et al*., 2004b, Klein, Kotov and Bufferd, 2011, Köhler *et al*., 2018, Santini *et al*., 2016, Vinkers *et al*., 2014, Zhou *et al*., 2019) and tested if they were associated with both depression and TLEs (data not shown). Sex, marital status and working status (Kaprio and Koskenvuo, 1988, Koskenvuo, Langinvainio, Kaprio and Sarna, 1984), emotional support (Dalgard, Bjørk and Tambs, 1995, Romanov, Varjonen, Kaprio and Koskenvuo, 2003) and ACE (Carlson *et al*., 2011) were used as sum or categorical/dichotomous variables (any vs. none). Age, formal education years until 1981 (Silventoinen, Krueger, Bouchard, Kaprio and McGue, 2004), a 10-item neuroticism scale (Floderus, 1974, Rose, Koskenvuo, Kaprio, Sarna and Langinvainio, 1988), and time-weighted (during the last six months [highest points], during the last 5 yrs, earlier, never) sum of 12 negative life events (death of a spouse, close relative or a friend, individual health or sexual problems, family member’s poor health, substantial conflicts, divorce or legal separation with a spouse, problems in other human relationships, considerable interpersonal conflicts at work, loss of job, serious financial problems, and other serious problem, range) (Romanov *et al*., 2003) were used as continuous variables. Formations of the variables are described in detail elsewhere (Piirtola *et al*., 2021, Romanov *et al*., 2003). Table 1 describes the covariates by sex and depression status.

### Statistical methods

#### Individual-based analyses

Depression sum score could be calculated for 97% (n=8,165) of the cohort (Figure 1). Prevalence of depression and TLEs were calculated for the total cohort and separately for men and women. Difference between sexes was analysed using chi-squared tests.

Odds Ratios (OR) with 95% Confidence Intervals (CI) from logistic regression models assessed the strength of associations between TLE and depression. Analyses were restricted to those individuals who had information about all covariates (n=6,002, Figure 1). Analyses were performed for each TLE separately at four stages. Model 1 was adjusted for age and sex. Model 2 was adjusted for age, sex, education, neuroticism, and emotional support. Model 3 was adjusted for covariates in model 2 plus other negative life events, whereas Model 4 was additionally adjusted for any ACE instead of negative life events, because we wanted to observe confounding effects of those 2 variables separately.

Regression analyses were performed first separately for men and women (Heidari, Babor, De Castro, Tort and Curno, 2016). Then interactions between sex and TLE, ACE and TLE, and ACE sum and TLE on depression were analysed with Likelihood-ratio test comparing two nested models with and without interaction term. If no sex interaction was found (p>0.05), analyses were performed by pooling men and women.

A drop-out analysis using age- and sex-adjusted logistic regression analyses related to depression was performed for those excluded from the main analyses (Figure 1, Table S1).

In all analyses of individuals, we corrected for the statistical dependence of twins within the twin pairs (Williams, 2000).

#### Within-pair analyses

Twin pairs generally share the same childhood environment and experiences. DZ pairs are genetically full siblings whereas MZ pairs are almost identical at the genomic sequence (Jonsson *et al*., 2021). Therefore, twin pairs discordant for depression are informative concerning the contribution of familial confounding (genetics and shared environmental) in the association between TLE and depression. In such pairs, one twin has depression and the other does not, and the analysis examines the distribution of TLE in such pairs, using principles and approaches described earlier (Piirtola *et al*., 2021, van Dongen, Slagboom, Draisma, Martin and Boomsma, 2012). Results from the within-pair analyses are compared with the results from individual-based analyses. If familial confounding plays a significant role in the association between TLE and depression, we should see an association among all individuals but not within twin pairs. Examining MZ and DZ pairs separately assesses whether familial confounding is due to shared environment or genetic influences.

Within-pair analyses were done using conditional logistic regression by zygosity in twin pairs discordant for depression (Figure 1b), first for all pairs, and then separately by zygosity. We tested for heterogeneity by zygosity using the matched-pair cohort method (csmatch test in Stata)(Cummings and McKnight, 2004). Within-pair analyses were performed first separately for men and women. If no sex differences in the OR estimates were found, men and women were analysed together. Notably, in addition to age, sex is automatically controlled for in the same-sexed twin pairs.

Stata SE version 18 (StataCorp, College Station, Texas, USA) was used for all the analyses. We report two-tailed p-values, unless otherwise stated.

## RESULTS

### Individual-based analyses

The prevalence of depression in the whole cohort was 13.3%, being higher among women (15.0%) compared with men (11.2%), p<0.001 (Table 1). Distributions of the CES-D scale sum in the whole cohort and by sex are shown in the Supplement Figure 1.

Of the 7,250 participants providing information about TLEs, most (70.8%) reported no TLEs in adulthood (men 69.4%; women 71.9%). One or two different kinds of TLEs were reported by 27.5% (men 29.1%; women 26.3%) and at least three TLEs by 1.7% (1.6% men; 1.8% women) of the participants. The mean sum of TLEs in adulthood was 0.40 (SD 0.71, range 0-6) in men and 0.38 (SD 0.70, range 0-4) in women. Men reported more involvements in serious traffic accidents and other serious accidents and being a victim of or witness to a violent crime than women (Table 2). Women reported 10 times more often sexual assault than men. There were no other major differences in the exposure to the reported TLEs between sexes.

Depression was generally more common among those men and women reporting TLEs, but the frequencies varied greatly across the TLEs (Table 2). In the age- and sex-adjusted analyses (Model 1), we found evidence of associations of serious accidents (OR 1.36; 95% CI 1.01, 1.85), physical assault (OR 3.10; 95% CI 2.45, 3.93), sexual assault (OR 3.49; 95% CI 2.67, 4.55), and violent crime (OR 3.86; 95% CI 2.59, 5.73) with depression (Table 3). The associations of physical assault, sexual assault, and violent crime remained substantial with no attenuation of the strength of association in any of the adjusted models (Models 2 – 4). Overall, we did not find evidence for sex interactions (i.e., sex differences in the OR estimates) (Table S2).

**Table 3.**
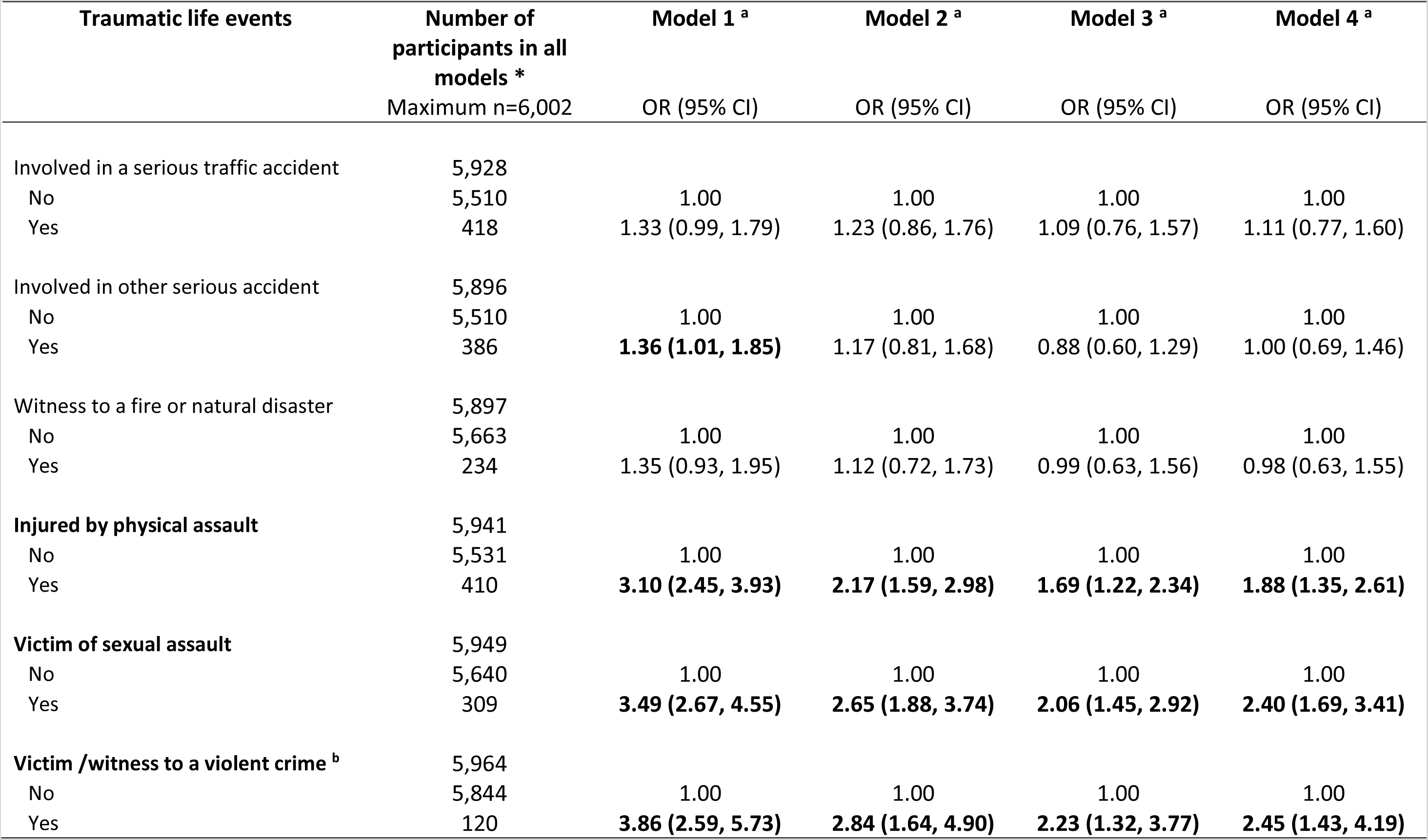
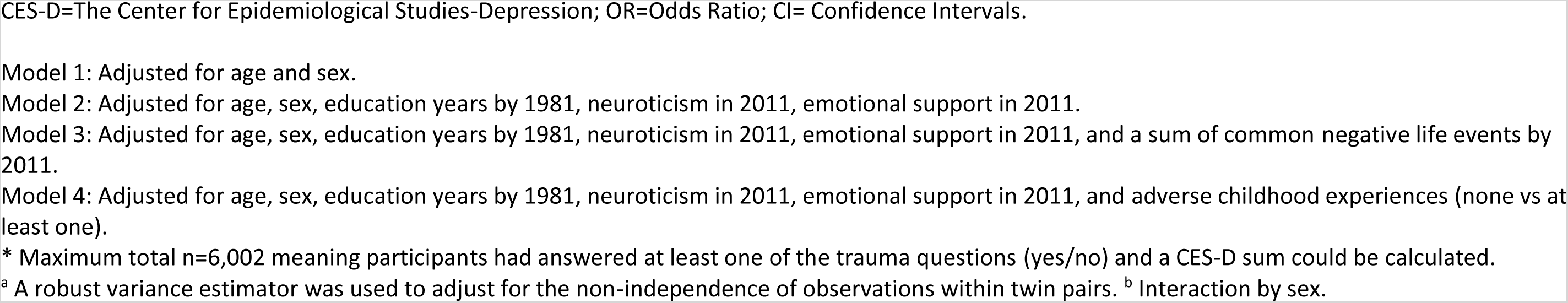
Associations (Odds Ratio [OR] with 95% Confidence Intervals [CI]) of adulthood traumatic life events with depression (The Center for Epidemiological Studies-Depression [CES-D] ≥20) in 6,002 participants of the Finnish Twin Cohort in 2011.

| Traumatic life events | Number of participants in all models * | Model 1 <sup>a</sup> | Model 2 <sup>a</sup> | Model 3 <sup>a</sup> | Model 4 <sup>a</sup> |
| --- | --- | --- | --- | --- | --- |
|  | Maximum n=6,002 | OR (95% CI) | OR (95% CI) | OR (95% CI) | OR (95% CI) |
| Involved in a serious traffic accident | 5,928 |  |  |  |  |
| No | 5,510 | 1.00 | 1.00 | 1.00 | 1.00 |
| Yes | 418 | 1.33 (0.99, 1.79) | 1.23 (0.86, 1.76) | 1.09 (0.76, 1.57) | 1.11 (0.77, 1.60) |
| Involved in other serious accident | 5,896 |  |  |  |  |
| No | 5,510 | 1.00 | 1.00 | 1.00 | 1.00 |
| Yes | 386 | **1.36 (1.01, 1.85)** | 1.17 (0.81, 1.68) | 0.88 (0.60, 1.29) | 1.00 (0.69, 1.46) |
| Witness to a fire or natural disaster | 5,897 |  |  |  |  |
| No | 5,663 | 1.00 | 1.00 | 1.00 | 1.00 |
| Yes | 234 | 1.35 (0.93, 1.95) | 1.12 (0.72, 1.73) | 0.99 (0.63, 1.56) | 0.98 (0.63, 1.55) |
| Injured by physical assault | 5,941 |  |  |  |  |
| No | 5,531 | 1.00 | 1.00 | 1.00 | 1.00 |
| Yes | 410 | **3.10 (2.45, 3.93)** | **2.17 (1.59, 2.98)** | **1.69 (1.22, 2.34)** | **1.88 (1.35, 2.61)** |
| Victim of sexual assault | 5,949 |  |  |  |  |
| No | 5,640 | 1.00 | 1.00 | 1.00 | 1.00 |
| Yes | 309 | **3.49 (2.67, 4.55)** | **2.65 (1.88, 3.74)** | **2.06 (1.45, 2.92)** | **2.40 (1.69, 3.41)** |
| Victim /witness to a violent crime <sup>b</sup> | 5,964 |  |  |  |  |
| No | 5,844 | 1.00 | 1.00 | 1.00 | 1.00 |
| Yes | 120 | **3.86 (2.59, 5.73)** | **2.84 (1.64, 4.90)** | **2.23 (1.32, 3.77)** | **2.45 (1.43, 4.19)** |

CES-D=The Center for Epidemiological Studies-Depression; OR=Odds Ratio; CI= Confidence Intervals.
Model 1: Adjusted for age and sex.
Model 2: Adjusted for age, sex, education years by 1981, neuroticism in 2011, emotional support in 2011.
Model 3: Adjusted for age, sex, education years by 1981, neuroticism in 2011, emotional support in 2011, and a sum of common negative life events by 2011.
Model 4: Adjusted for age, sex, education years by 1981, neuroticism in 2011, emotional support in 2011, and adverse childhood experiences (none vs at least one).
\* Maximum total n=6,002 meaning participants had answered at least one of the trauma questions (yes/no) and a CES-D sum could be calculated.
<sup>a</sup> A robust variance estimator was used to adjust for the non-independence of observations within twin pairs. <sup>b</sup> Interaction by sex.

The likelihood for depression was higher among those reporting more TLEs. In age- and sex-adjusted analyses, those reporting one to two TLEs had about 2-times higher likelihood for depression (OR 1.92; 95% CI 1.61, 2.29) than those with no TLEs, and the odd ratios were 4-fold for those reporting at least three TLEs (OR 4.54; 95% CI 2.83, 7.28). There was no statistical interaction between the number of TLEs and sex on depression. In the multivariate adjusted analyses (models 2-4), the strength of odds ratios attenuated slightly in one/two TLE category on depression (ORs varied from 1.23 to 1.52) but among those with at least three TLEs the association with depression attenuated (ORs varied from 2.29 to 3.26) (detailed data not shown).

A statistical interaction (p=0.018) was found between the numbers of TLEs and of ACEs on depression. Among specific TLEs, a statistically significant interaction was found only between sum on ACEs and involvement in other serious accidents (p=0.006) but not between any other TLEs. Further, interaction between ACE sum and involvement in other serious accidents was statistically significant only in men (p=0.036).

Results from the drop-out analysis related to depression are shown in Table S1.

### Within-pair analyses

Altogether, there were 399 pairs discordant for depression (264 DZ, 135 MZ) (Fig 1b). The within-pairs associations of physical assault and sexual assault were elevated and statistically significant in all pairs, DZ pairs and MZ pairs (Table 4). The associations of violent crime with depression were of the same strength across the three groups of pairs, though not significant for MZ pairs. Associations of involvement in other serious accidents with depression was seen only in DZ pairs. Overall, within-pair associations between TLEs and depression did not differ by sex (Table S3).

**Table 4.** Within-pair associations (Odds Ratio [OR] with 95% Confidence Intervals [CI]) of adulthood traumatic life events with depression (The Center for Epidemiological Studies-Depression [CES-D] ≥20) in pairs discordant for depression. All pairs and by zygosity in the Finnish Twin Cohort in 2011.

| Traumatic life events | ALL | DZ pairs | MZ pairs | Homogeneity test between DZ and MZ pairs |
| --- | --- | --- | --- | --- |
|  | N<br>OR<br>(95% CI) | N<br>OR<br>(95% CI) | N<br>OR<br>(95% CI) | p-value |
| Involved in a serious traffic accident | 377<br>1.10<br>(0.67, 1.78) | 252<br>1.22<br>(0.66, 2.28) | 125<br>0.92<br>(0.42, 2.02) | 0.564 |
| Involved in other serious accident | 367<br>1.56<br>(0.94, 2.58) | 246<br>**1.94**<br>**(1.06, 3.54)** | 121<br>0.89<br>(0.34, 2.30) | 0.252 |
| Witness to a fire or natural disaster | 373<br>1.12<br>(0.58, 2.15) | 252<br>0.86<br>(0.40, 1.85) | 121<br>2.33<br>(0.60, 9.02) | 0.195 |
| Injured by physical assault | 380<br>**2.90**<br>**(1.77, 4.77)** | 252<br>**2.85**<br>**(1.51, 5.35)** | 128<br>**3.00**<br>**(1.35, 6.68)** | 0.596 |
| Victim of sexual assault | 378<br>**2.89**<br>**(1.69, 4.94)** | 251<br>**2.92**<br>**(1.51, 5.62)** | 127<br>**2.83**<br>**(1.12, 7.19)** | 1.00 |
| Victim of or witness to a violent crime | 383<br>**3.00**<br>**(1.35, 6.68)** | 255<br>**2.67**<br>**(1.04, 6.81)** | 128<br>4.00<br>(0.85, 18.8) | 0.460 |

CES-D=The Center for Epidemiological Studies-Depression; OR=Odds Ratio measured with conditional logistic regression analysis; CI= Confidence Intervals; N=number of pairs.

## DISCUSSION

In this study, we examined the sex-based prevalence of adulthood traumatic life events and analysed whether the associations of TLEs with depression were independent of vulnerability, exposure loading, measured covariates or familial confounding. We found support for sex-based trauma exposure differences but not for sex-based vulnerability for depression after trauma experience. Men were exposed more often to traffic accidents, other serious accidents, and violent crime than women, whereas women reported ten times more sexual assault than men. Thus, the exposure to various TLEs seems to differ by sex which, at least partly, supports the exposure hypothesis. However, some traumas were more strongly associated with depression than others. In our study, serious accidents, physical assault, sexual assault, and violent crime were all associated with depression whereas being involved in a serious traffic accident, or in other serious accidents, or witnessing fires or natural disasters were not associated with depression. The associations were robust to adjustment for individual covariates or for shared familial factors. Further, even though depression was more common among women compared with men, we did not find evidence that associations between TLEs and depression were modified by sex. Therefore, our results implicate that if exposed to TLEs, both men and women have similar vulnerability for depression. The associations appear to be independent of several individual covariates, including ACEs and neuroticism, as well of shared familial factors including genetic vulnerability.

We observed that the prevalence of depression was higher in women than in men. This sex ratio is consistent with most earlier studies (Bebbington, 1996, Bromet *et al*., 2011, Markkula *et al*., 2015, Salk *et al*., 2017, Steel *et al*., 2014).

Most (70%) people worldwide report exposure to at least one TLE during their lifespan (Benjet *et al*., 2016, Kessler *et al*., 2017). In our study, the proportion was 32% in adulthood. Notably, there is great variation in TLE prevalence across countries (Benjet *et al*., 2016), which may be partly accounted for by which TLEs are assessed. In addition, social unrest, conflicts, and natural catastrophes, affect some countries more than others. We included only a relatively small number of serious TLEs in our analyses, while other adverse life events were categorized as negative life events. When we combined lifelong TLEs with negative life events, their combined prevalence was 78%. Globally, the most common TLEs have been events that cause bodily harm for someone (Benjet *et al*., 2016). This finding is in line with our results.

In line with the exposure hypothesis (Denton *et al*., 2004), we observed sex-based differences in exposure to traumas. Men were more exposed to traffic accidents, other serious accidents, and violent crime than women, who were more exposed to sexual assault. Exposure to domestic violence and sexual assault has been reported to be higher in females than in males (Attila *et al*., 2023, Fergusson, Swain-Campbell and Horwood, 2002). In previous studies, men reported more TLE categories overall than women during the life span (Hatch and Dohrenwend, 2007), but we did not observe this for either TLEs in adulthood or ACEs in childhood. Altogether, men and women seem to be exposed equally to traumatic and adverse life events, but the type of event varies.

In a meta-analysis, over half of individuals with current post-traumatic stress disorder had also major depressive disorder (Rytwinski, Scur, Feeny and Youngstrom, 2013). In our study, both the cumulative load of TLEs and some specific TLEs were associated with depression, as observed earlier (Kessler, 1997, McCutcheon *et al*., 2009, Wang *et al*., 2023, Xiang and Wang, 2021). We found that three traumas were associated with depression among both men and women: being injured by physical assault, being victim of sexual assault and being victim or witnessing to a violent crime. The associations remained substantial with no attenuation of the strength of association in all adjusted models.

A high prevalence of sexual assault has been reported in many countries (Kessler *et al*., 2017). We found that sexual assault was reported by ten times more women than men. Nonetheless, the association with depression was of the same magnitude in men and women and was independent of measured covariates. In the within-pair analyses, shared exposures and experiences between twin siblings did not attenuate the association, nor did control of genetic relatedness. Thus, possible shared genetic liability between TLEs and depression does not account for the association. Indeed, we provide further evidence for a causal path from sexual assault to depression.

In addition to sexual assault, being injured by physical assault or being victim or witnessing to a violent crime can be categorized as type II traumas (Wang *et al*., 2023). Because these specific traumas were associated with depression both in men and women, our findings do not support gender vulnerability hypothesis for depression due these traumas (Bilodeau *et al*., 2020, Piccinelli and Wilkinson, 2000). The individual-based modelling of covariates and within-pair analyses are consistent with a causal role of violent crime victimization in the development of depression.

We further considered cumulative load of ACEs, TLEs and weighted sum of other negative life events during the adulthood in our analyses, but they did not substantially impact the association of adult TLEs and depression. However, we found statistically significant sex-interaction on depression between sum on ACEs and involvement in other serious accidents in adulthood, yet only in males. In previous studies, individuals who have experienced ACEs or TLEs, have been reported to be at an increased risk of further TLEs and this cumulative chain of adversities can affect individual’s later well-being and depression risk (Bürgin *et al*., 2021, Kendler *et al*., 2004b, McLaughlin *et al*., 2010, Pearlin *et al*., 2005). Also, a dose-response relationship between stressors and depression has been shown indicating that higher amount or ACEs or TLS in adulthood lower the threshold for depression (Bandoli *et al*., 2017, Kendler *et al*., 2004b, McLaughlin *et al*., 2010).

Personality traits and emotional dysregulation have been proposed as important modifiers between men and women for the onset of depression explaining some part of prevalence differences of depression between genders (Parker and Brotchie, 2010). Women with high neuroticism scores have reported health problems through different kinds of TLEs (Iacovino, Bogdan and Oltmanns, 2016). In our analyses, including neuroticism in the model weakened modestly the association of depression with all kinds of traumas, however, with a statistically significant effect only for involvement in other serious accidents. Thus, our analyses slightly support previous findings about the role of personality in the development of depression after stressful life events (Iacovino *et al*., 2016).

Even though TLEs increase the risk of major depression in later life (McCutcheon *et al*., 2009, Vibhakar *et al*., 2019, Xiang and Wang, 2021), not all individuals develop depressive disorders, and multiple protective factors and pathways to resilience exist (Bonanno, 2004, Smith and Hayslip Jr, 2012). We did not test the resilience hypothesis as such, but we adjusted our analyses for several factors related to resilience in relation to depressive disorders.

### Strengths

Our data is a relatively large population-based sample with high participation rate including both men and women. We have information about multiple potential covariates. Uniquely, we could take account familial factors including genetics.

### Limitations

Our data came from a survey where depressive symptoms were reported cross-sectionally using the CES-D scale, and we have not compared it to diagnostic measures of major depressive disorder. However, a sensitivity of 0.83 and specificity of 0.78 for individuals with clinically relevant level of depression has been shown with a cut off value ≥20 (Vilagut *et al*., 2016). However, reverse causation cannot rule out as a limitation. The age group in this study represents late adulthood (mean age 60) and there may be some bias if those with severe depression have not replied.

The TLEs were reported retrospectively. However, retrospective reports are shown to be useful risk indicators of TLEs associated with incidence of psychopathology and its course of illness (Baldwin *et al*., 2019). Depressive disorders can increase reporting previous traumatic life event (Baldwin *et al*., 2019, Schraedley, Turner and Gotlib, 2002). On the other hand, both depressive individuals and those who experienced TLEs might be missing from the analyses. We do not know whether respondents were not answering some TLE items because they have not experienced any TLEs or whether they found it too intimate or painful to answer. We chose a relatively small set of relevant TLEs from the Trauma History Screen measure (Carlson *et al*., 2011), while other studies have assessed more traumas and adverse life events (Kessler, 1997, Kessler *et al*., 2017, McCutcheon *et al*., 2009, Vibhakar *et al*., 2019, Wang *et al*., 2023, Xiang and Wang, 2021). We did analyse the effect of cumulative sum of TLEs on depression, but we have no information when or in which order TLEs have happened to a person during adulthood and whether someone has experienced several events of the same nature.

### Further studies

Even though there is evidence that TLEs increase the risk of major depression in later life (McCutcheon *et al*., 2009, Vibhakar *et al*., 2019, Xiang and Wang, 2021), not all individuals develop depressive disorders due to multiple protective factors (Bonanno, 2004, Smith and Hayslip Jr, 2012). We controlled for many resilience related factors like emotional support, but such adjustments did not substantially reduce the odds for depression especially among those injured by physical assault or being victims of sexual assault. The role of resilience related factors after a trauma as well as the type and order of cumulative load of traumatic events during lifespan warrants more studies.

## CONCLUSIONS

Women seem to suffer from depression more than men during middle-age. Men are exposed more to traffic accidents and other serious accidents, as well as to violent crime, whereas women are exposed more to sexual assault. Traumatic life events are robustly associated with depression independently of sex and these associations appear to be independent of the variables increasing vulnerability for depression, such as personality, socioeconomic factors, other negative life events, and ACEs, as well as of familial influences, including shared genetic liability. Exposure to traumatic life events should be addressed in prevention and treatment of depression.

## Data Availability

All data produced in the present study are available upon reasonable request to the authors

## Acknowledgements

We acknowledge all twin participants for their valuable co-operation along the years.

## REFERENCES

Attila, H., Keski-Petäjä, M., Pietiläinen, M., Lipasti, L., Saari, J. & Haapakangas, K. (2023). Sukupuolistunut väkivalta ja lähisuhdeväkivalta Suomessa 2021. Loppuraportti (Gendered violence and domestic violence in Finland 2021. Final report). Tilastokeskus (https://urn.fi/URN:ISBN:978-952-244-717-3, 18.2.2024)

Baldwin, J. R., Reuben, A., Newbury, J. B. & Danese, A. (2019). Agreement Between Prospective and Retrospective Measures of Childhood Maltreatment: A Systematic Review and Meta-analysis. JAMA psychiatry (Chicago, Ill.) 76, 584–593.

Bandoli, G., Campbell-Sills, L., Kessler, R. C., Heeringa, S. G., Nock, M. K., Rosellini, A. J., Sampson, N. A., Schoenbaum, M., Ursano, R. J. & Stein, M. B. (2017). Childhood adversity, adult stress, and the risk of major depression or generalized anxiety disorder in US soldiers: a test of the stress sensitization hypothesis. Psychological Medicine 47, 2379–2392.

Bebbington, P. (1996). The origins of sex differences in depressive disorder: bridging the gap. International Review of Psychiatry 8, 295–332.

Benjet, C., Bromet, E., Karam, E. G., Kessler, R. C., McLaughlin, K. A., Ruscio, A. M., Shahly, V., Stein, D. J., Petukhova, M., Hill, E., Alonso, J., Atwoli, L., Bunting, B., Bruffaerts, R., Caldas-de-Almeida, J. M., de Girolamo, G., Florescu, S., Gureje, O., Huang, Y., Lepine, J. P., Kawakami, N., Kovess-Masfety, V., Medina-Mora, M. E., Navarro-Mateu, F., Piazza, M., Posada-Villa, J., Scott, K. M., Shalev, A., Slade, T., ten Have, M., Torres, Y., Viana, M. C., Zarkov, Z. & Koenen, K. C. (2016). The epidemiology of traumatic event exposure worldwide: results from the World Mental Health Survey Consortium. Psychol Med 46, 327–43.

Bilodeau, J., Marchand, A. & Demers, A. (2020). Work, family, work-family conflict and psychological distress: A revisited look at the gendered vulnerability pathways. Stress Health 36, 75–87.

Bonanno, G. A. (2004). Loss, trauma, and human resilience: have we underestimated the human capacity to thrive after extremely aversive events? Am Psychol 59, 20–8.

Bromet, E., Andrade, L. H., Hwang, I., Sampson, N. A., Alonso, J., de Girolamo, G., de Graaf, R., Demyttenaere, K., Hu, C., Iwata, N., Karam, A. N., Kaur, J., Kostyuchenko, S., Lepine, J. P., Levinson, D., Matschinger, H., Mora, M. E., Browne, M. O., Posada-Villa, J., Viana, M. C., Williams, D. R. & Kessler, R. C. (2011). Cross-national epidemiology of DSM-IV major depressive episode. BMC Med 9, 90.

Bürgin, D., Boonmann, C., Schmeck, K., Schmid, M., Tripp, P., Nishimi, K. & O’Donovan, A. (2021). Compounding Stress: Childhood Adversity as a Risk Factor for Adulthood Trauma Exposure in the Health and Retirement Study. Journal of traumatic stress 34, 124–136.

Carlson, E. B., Smith, S. R., Palmieri, P. A., Dalenberg, C., Ruzek, J. I., Kimerling, R., Burling, T. A. & Spain, D. A. (2011). Development and validation of a brief self-report measure of trauma exposure: the Trauma History Screen. Psychol Assess 23, 463–77.

Cummings, P. & McKnight, B. (2004). Analysis of matched cohort data. Stata Journal 4, 274–281.

Dalgard, O. S., Bjørk, S. & Tambs, K. (1995). Social support, negative life events and mental health. Br J Psychiatry 166, 29–34.

Denton, M., Prus, S. & Walters, V. (2004). Gender differences in health: a Canadian study of the psychosocial, structural and behavioural determinants of health. Soc Sci Med 58, 2585–600.

Fergusson, D. M., Swain-Campbell, N. R. & Horwood, L. J. (2002). Does sexual violence contribute to elevated rates of anxiety and depression in females? Psychol Med 32, 991–6.

Flint, J. & Kendler, Kenneth S. (2014). The Genetics of Major Depression. Neuron 81, 484–503.

Floderus, B. (1974). Psycho-social factors in relation to coronary heart disease and associated risk factors. Nordisk Hygienisk Tidskrift Suppl 6, 7–148.

Hammen, C., Henry, R. & Daley, S. E. (2000). Depression and sensitization to stressors among young women as a function of childhood adversity. J Consult Clin Psychol 68, 782–7.

Hatch, S. L. & Dohrenwend, B. P. (2007). Distribution of traumatic and other stressful life events by race/ethnicity, gender, SES and age: a review of the research. Am J Community Psychol 40, 313–32.

Heidari, S., Babor, T. F., De Castro, P., Tort, S. & Curno, M. (2016). Sex and Gender Equity in Research: rationale for the SAGER guidelines and recommended use. Research Integrity and Peer Review 1, 2.

Hyde, J. S., Mezulis, A. H. & Abramson, L. Y. (2008). The ABCs of depression: integrating affective, biological, and cognitive models to explain the emergence of the gender difference in depression. Psychol Rev 115, 291–313.

Iacovino, J. M., Bogdan, R. & Oltmanns, T. F. (2016). Personality Predicts Health Declines Through Stressful Life Events During Late Mid-Life. Journal of Personality 84, 536–546.

Jonsson, H., Magnusdottir, E., Eggertsson, H. P., Stefansson, O. A., Arnadottir, G. A., Eiriksson, O., Zink, F., Helgason, E. A., Jonsdottir, I., Gylfason, A., Jonasdottir, A., Jonasdottir, A., Beyter, D., Steingrimsdottir, T., Norddahl, G. L., Magnusson, O. T., Masson, G., Halldorsson, B. V., Thorsteinsdottir, U., Helgason, A., Sulem, P., Gudbjartsson, D. F. & Stefansson, K. (2021). Differences between germline genomes of monozygotic twins. Nat Genet 53, 27–34.

Kaprio, J. (2013). The Finnish Twin Cohort Study: an update. Twin Res Hum Genet 16, 157–62.

Kaprio, J. & Koskenvuo, M. (1988). A prospective study of psychological and socioeconomic characteristics, health behavior and morbidity in cigarette smokers prior to quitting compared to persistent smokers and non-smokers. J Clin Epidemiol 41, 139–50.

Kendall, K. M., Van Assche, E., Andlauer, T. F. M., Choi, K. W., Luykx, J. J., Schulte, E. C. & Lu, Y. (2021). The genetic basis of major depression. Psychological Medicine, 1–14.

Kendler, K. S. & Gardner, C. O. (2014). Sex differences in the pathways to major depression: a study of opposite-sex twin pairs. Am J Psychiatry 171, 426–35.

Kendler, K. S., Kuhn, J. & Prescott, C. A. (2004a). The interrelationship of neuroticism, sex, and stressful life events in the prediction of episodes of major depression. Am J Psychiatry 161, 631–6.

Kendler, K. S., Kuhn, J. W. & Prescott, C. A. (2004b). Childhood sexual abuse, stressful life events and risk for major depression in women. Psychol Med 34, 1475–82.

Kendler, K. S., Thornton, L. M. & Prescott, C. A. (2001). Gender differences in the rates of exposure to stressful life events and sensitivity to their depressogenic effects. Am J Psychiatry 158, 587–93.

Kessler, R. C. (1997). The effects of stressful life events on depression. Annual review of psychology 48, 191–214.

Kessler, R. C., Aguilar-Gaxiola, S., Alonso, J., Benjet, C., Bromet, E. J., Cardoso, G., Degenhardt, L., de Girolamo, G., Dinolova, R. V., Ferry, F., Florescu, S., Gureje, O., Haro, J. M., Huang, Y., Karam, E. G., Kawakami, N., Lee, S., Lepine, J. P., Levinson, D., Navarro-Mateu, F., Pennell, B. E., Piazza, M., Posada-Villa, J., Scott, K. M., Stein, D. J., Ten Have, M., Torres, Y., Viana, M. C., Petukhova, M. V., Sampson, N. A., Zaslavsky, A. M. & Koenen, K. C. (2017). Trauma and PTSD in the WHO World Mental Health Surveys. Eur J Psychotraumatol 8, 1353383.

Klein, D. N., Kotov, R. & Bufferd, S. J. (2011). Personality and depression: explanatory models and review of the evidence. Annual review of clinical psychology 7, 269–295.

Koskenvuo, M., Langinvainio, H., Kaprio, J. & Sarna, S. (1984). Health related psychosocial correlates of neuroticism: a study of adult male twins in Finland. Acta Genet Med Gemellol (Roma) 33, 307–20.

Köhler, C. A., Evangelou, E., Stubbs, B., Solmi, M., Veronese, N., Belbasis, L., Bortolato, B., Melo, M. C. A., Coelho, C. A., Fernandes, B. S., Olfson, M., Ioannidis, J. P. A. & Carvalho, A. F. (2018). Mapping risk factors for depression across the lifespan: An umbrella review of evidence from meta-analyses and Mendelian randomization studies. J Psychiatr Res 103, 189–207.

Luo, J., Zhang, B. & Roberts, B. W. (2021). Sensitization or inoculation: Investigating the effects of early adversity on personality traits and stress experiences in adulthood. PloS one 16, e0248822–e0248822.

Lyons, M. J., Goldberg, J., Eisen, S. A., True, W., Tsuang, M. T., Meyer, J. M. & Henderson, W. G. (1993). Do genes influence exposure to trauma? A twin study of combat. Am J Med Genet 48, 22–7.

Maji, S. (2018). Society and ‘good woman’: A critical review of gender difference in depression. Int J Soc Psychiatry 64, 396–405.

Markkula, N., Suvisaari, J., Saarni, S. I., Pirkola, S., Pena, S., Saarni, S., Ahola, K., Mattila, A. K., Viertio, S., Strehle, J., Koskinen, S. & Harkanen, T. (2015). Prevalence and correlates of major depressive disorder and dysthymia in an eleven-year follow-up--results from the Finnish Health 2011 Survey. J Affect Disord 173, 73–80.

McCutcheon, V. V., Heath, A. C., Nelson, E. C., Bucholz, K. K., Madden, P. A. & Martin, N. G. (2009). Accumulation of trauma over time and risk for depression in a twin sample. Psychol Med 39, 431–41.

McGue, M., Osler, M. & Christensen, K. (2010). Causal Inference and Observational Research: The Utility of Twins. Perspect Psychol Sci 5, 546–556.

McLaughlin, K. A., Conron, K. J., Koenen, K. C. & Gilman, S. E. (2010). Childhood adversity, adult stressful life events, and risk of past-year psychiatric disorder: a test of the stress sensitization hypothesis in a population-based sample of adults. Psychological medicine 40, 1647–1658.

Otten, D., Tibubos, A. N., Schomerus, G., Brähler, E., Binder, H., Kruse, J., Ladwig, K. H., Wild, P. S., Grabe, H. J. & Beutel, M. E. (2021). Similarities and Differences of Mental Health in Women and Men: A Systematic Review of Findings in Three Large German Cohorts. Front Public Health 9, 553071.

Parker, G. & Brotchie, H. (2010). Gender differences in depression. International Review of Psychiatry 22, 429–436.

Pearlin, L. I., Schieman, S., Fazio, E. M. & Meersman, S. C. (2005). Stress, Health, and the Life Course: Some Conceptual Perspectives. Journal of health and social behavior 46, 205–219.

Piccinelli, M. & Wilkinson, G. (2000). Gender differences in depression: Critical review. British Journal of Psychiatry 177, 486–492.

Piirtola, M., Kaprio, J., Baker, T. B., Piasecki, T. M., Piper, M. E. & Korhonen, T. (2021). The associations of smoking dependence motives with depression among daily smokers. Addiction 116, 2162–2174.

Polderman, T. J., Benyamin, B., de Leeuw, C. A., Sullivan, P. F., van Bochoven, A., Visscher, P. M. & Posthuma, D. (2015). Meta-analysis of the heritability of human traits based on fifty years of twin studies. Nat Genet.

Power, R. A., Wingenbach, T., Cohen-Woods, S., Uher, R., Ng, M. Y., Butler, A. W., Ising, M., Craddock, N., Owen, M. J., Korszun, A., Jones, L., Jones, I., Gill, M., Rice, J. P., Maier, W., Zobel, A., Mors, O., Placentino, A., Rietschel, M., Lucae, S., Holsboer, F., Binder, E. B., Keers, R., Tozzi, F., Muglia, P., Breen, G., Craig, I. W., Muller-Myhsok, B., Kennedy, J. L., Strauss, J., Vincent, J. B., Lewis, C. M., Farmer, A. E. & McGuffin, P. (2013). Estimating the heritability of reporting stressful life events captured by common genetic variants. Psychol Med 43, 1965–71.

Radloff, L. S. (1977). The CES-D Scale:A Self-Report Depression Scale for Research in the General Population. Applied Psychological Measurement 1, 385–401.

Richardson, S., Carr, E., Netuveli, G. & Sacker, A. (2020). Adverse events over the life course and later-life wellbeing and depressive symptoms in older people. Int Psychogeriatr, 1–15.

Romanov, K., Varjonen, J., Kaprio, J. & Koskenvuo, M. (2003). Life events and depressiveness - the effect of adjustment for psychosocial factors, somatic health and genetic liability. Acta Psychiatr Scand 107, 25–33.

Rose, R. J., Koskenvuo, M., Kaprio, J., Sarna, S. & Langinvainio, H. (1988). Shared genes, shared experiences, and similarity of personality: data from 14,288 adult Finnish co-twins. J Pers Soc Psychol 54, 161–71.

Rytwinski, N. K., Scur, M. D., Feeny, N. C. & Youngstrom, E. A. (2013). The co-occurrence of major depressive disorder among individuals with posttraumatic stress disorder: a meta-analysis. J Trauma Stress 26, 299–309.

Salk, R. H., Hyde, J. S. & Abramson, L. Y. (2017). Gender differences in depression in representative national samples: Meta-analyses of diagnoses and symptoms. Psychol Bull 143, 783–822.

Santini, Z. I., Fiori, K. L., Feeney, J., Tyrovolas, S., Haro, J. M. & Koyanagi, A. (2016). Social relationships, loneliness, and mental health among older men and women in Ireland: A prospective community-based study. Journal of affective disorders 204, 59–69.

Schraedley, P. K., Turner, R. J. & Gotlib, I. H. (2002). Stability of retrospective reports in depression: traumatic events, past depressive episodes, and parental psychopathology. J Health Soc Behav 43, 307–16.

Silventoinen, K., Krueger, R. F., Bouchard, T. J., Jr., Kaprio, J. & McGue, M. (2004). Heritability of body height and educational attainment in an international context: comparison of adult twins in Minnesota and Finland. Am J Hum Biol 16, 544–55.

Smith, G. C. & Hayslip Jr, B. (2012). Chapter 1. Resilience in Adulthood and Later Life: What Does it Mean and Where Are We Heading? Annual Review of Gerontology and Geriatrics 32, 1–28.

Steel, Z., Marnane, C., Iranpour, C., Chey, T., Jackson, J. W., Patel, V. & Silove, D. (2014). The global prevalence of common mental disorders: a systematic review and meta-analysis 1980-2013. Int J Epidemiol 43, 476–93.

van Dongen, J., Slagboom, P. E., Draisma, H. H., Martin, N. G. & Boomsma, D. I. (2012). The continuing value of twin studies in the omics era. Nat Rev Genet 13, 640–53.

Vibhakar, V., Allen, L. R., Gee, B. & Meiser-Stedman, R. (2019). A systematic review and meta-analysis on the prevalence of depression in children and adolescents after exposure to trauma. J Affect Disord 255, 77–89.

Vilagut, G., Forero, C. G., Barbaglia, G. & Alonso, J. (2016). Screening for Depression in the General Population with the Center for Epidemiologic Studies Depression (CES-D): A Systematic Review with Meta-Analysis. PLoS One 11, e0155431.

Vinkers, C. H., Joëls, M., Milaneschi, Y., Kahn, R. S., Penninx, B. W. J. H. & Boks, M. P. M. (2014). STRESS EXPOSURE ACROSS THE LIFE SPAN CUMULATIVELY INCREASES DEPRESSION RISK AND IS MODERATED BY NEUROTICISM. Depression and anxiety 31, 737–745.

Wang, S. K., Feng, M., Fang, Y., Lv, L., Sun, G. L., Yang, S. L., Guo, P., Cheng, S. F., Qian, M. C. & Chen, H. X. (2023). Psychological trauma, posttraumatic stress disorder and trauma-related depression: A mini-review. World J Psychiatry 13, 331–339.

Williams, R. L. (2000). A note on robust variance estimation for cluster-correlated data. Biometrics 56, 645–6.

Xiang, X. & Wang, X. (2021). Childhood adversity and major depression in later life: A competing-risks regression analysis. International journal of geriatric psychiatry 36, 215–223.

Zhou, J., Feng, L., Hu, C., Pao, C., Xiao, L. & Wang, G. (2019). Associations Among Depressive Symptoms, Childhood Abuse, Neuroticism, Social Support, and Coping Style in the Population Covering General Adults, Depressed Patients, Bipolar Disorder Patients, and High Risk Population for Depression. Front Psychol 10, 1321.

Zimmermann, P., Brückl, T., Lieb, R., Nocon, A., Ising, M., Beesdo, K. & Wittchen, H. U. (2008). The interplay of familial depression liability and adverse events in predicting the first onset of depression during a 10-year follow-up. Biol Psychiatry 63, 406–14.

