## Supplemental materials for "Traumatic life events as predictors for depression in middle-aged men and women: A Finnish twin study"

**Other supplementary material**

**Table S1.** Associations of different groups with depression (The Center for Epidemiological Studies-Depression [CES-D] ≥20). Groups with or without information about traumas or covariates related to traumas compared to those included in the analyses (reference) (see Figure 1a).

| **Group** | **Number of individuals** | OR (95% CI) |
|---|---|---|
| Included in the individual-based analyses | 6,002 | reference |
| Excluded due to missing info of traumas | 1,047 | **1.41 (1.16, 1.70)** |
| Excluded due to missing info of covariates | 1,116 | **2.10 (1.78, 2.48)** |

CES-D=The Center for Epidemiological Studies-Depression. OR=Odds Ratio measured with logistic regression analysis; CI= Confidence Intervals. Statistically significant p-values (p<0.05) are in **bold**.

**Table S2.** Associations (Odds Ratio [OR] with 95% Confidence Intervals [CI]) of adulthood traumatic life events with depression (The Center for Epidemiological Studies-Depression [CES-D] ≥20) by sex in the Finnish Twin Cohort in 2011.

| **Traumatic life events** (number of events) | **Number of individuals in all models** n | **Model 1** [a] OR (95% CI) [b] | **Model 2** [a] OR (95% CI) [b] | **Model 3** [a] OR (95% CI) [b] | **Model 4** [a] OR (95% CI) [b] |
|---|---|---|---|---|---|
| **Involved in a serious traffic accident** | | | | | |
| Men (n=233) | 2,617 | 1.35 (0.88, 2.08) | 1.39 (0.82, 2.34) | 1.34 (0.80, 2.25) | 1.30 (0.77, 2.20) |
| Women (n=185) | 3,311 | 1.32 (0.88, 1.97) | 1.09 (0.69, 1.75) | 0.89 (0.55, 1.42) | 0.95 (0.57, 1.57) |
| LR test for sex difference (p-value) | | 0.942 | 0.464 | 0.242 | 0.465 |
| **Involved in other serious accident** | | | | | |
| Men (n=236) | 2,598 | **1.60 (1.07, 2.40)** | 1.34 (0.85, 2.11) | 1.09 (0.67, 1.76) | 1.25 (0.79, 2.00) |
| Women (n=150) | 3,298 | 1.13 (0.71, 1.80) | 0.97 (0.54, 1.76) | 0.69 (0.38, 1.25) | 0.74 (0.39, 1.39) |
| LR test for sex difference (p-value) | | 0.269 | 0.397 | 0.295 | 0.271 |
| **Witness to a fire or natural disaster** | | | | | |
| Men (n=103) | 2,598 | 1.09 (0.56, 2.11) | 0.87 (0.42, 1.81) | 0.76 (0.37, 1.57) | 0.77 (0.35, 1.66) |
| Women (n=131) | 3,299 | 1.51 (0.96, 2.38) | 1.30 (0.75, 2.26) | 1.18 (0.65, 2.14) | 1.15 (0.66, 2.01) |
| LR test for sex difference (p-value) | | 0.417 | 0.393 | 0.340 | 0.318 |
| **Injured by physical assault** | | | | | |
| Men (n=191) | 2,620 | **2.75 (1.88, 4.02)** | **1.68 (1.05, 2.69)** | 1.46 (0.90, 2.37) | 1.54 (0.95, 2.49) |
| Women (n=219) | 3,321 | **3.36 (2.48, 4.56)** | **2.74 (1.79, 4.21)** | **1.95 (1.26, 3.03)** | **2.30 (1.46, 3.59)** |
| LR test for sex difference (p-value) | | 0.443 | 0.137 | 0.363 | 0.180 |
| **Victim of sexual assault** | | | | | |
| Men (n=13) | 2,632 | 2.86 (0.75, 10.8) | 2.72 (0.36, 20.4) | 2.01 (0.22, 17.9) | 2.71 (0.33, 22.2) |
| Women (n=296) | 3,317 | **3.54 (2.70, 4.65)** | **2.70 (1.86, 3.91)** | **2.09 (1.43, 3.06)** | **2.40 (1.64, 3.51)** |
| LR test for sex difference (p-value) | | 0.776 | 0.939 | 0.996 | 0.853 |

(continues)

| **Traumatic life events** (number of events) | **Number of individuals in all models** n | **Model 1** [a] OR (95% CI) [b] | **Model 2** [a] OR (95% CI) [b] | **Model 3** [a] OR (95% CI) [b] | **Model 4** [a] OR (95% CI) [b] |
|---|---|---|---|---|---|
| **Victim of or witness to a violent crime** | | | | | |
| Men (n=63) | 2,637 | **2.89 (1.59, 5.25)** | 1.87 (0.90, 3.85) | 1.63 (0.82, 3.28) | 1.74 (0.85, 3.56) |
| Women (n=57) | 3,327 | **4.92 (2.83, 8.54)** | **4.42 (2.00, 9.89)** | **3.13 (1.40, 7.00)** | **3.58 (1.62, 7.92)** |
| LR test for sex difference (p-value) | | 0.204 | 0.120 | 0.234 | 0.157 |

CES-D=The Center for Epidemiological Studies-Depression; OR=Odds Ratio; CI= Confidence Intervals; LR test=Likelihood-ratio test for difference between sexes.
Model 1: Adjusted for age and sex.
Model 2: Adjusted for age, sex, education years by 1981, neuroticism in 2011, and emotional support in 2011.
Model 3: Adjusted for age, sex, education years by 1981, neuroticism in 2011, emotional support in 2011, and a sum of common negative life events by 2011.
Model 4: Adjusted for age, sex, education years by 1981, neuroticism in 2011, emotional support in 2011, and adverse childhood experiences (none vs at least one).
[a] A robust variance estimator was used to adjust for the non-independence of observations within twin pairs. [b] Reference group in Logistic regression model analyses those without a traumatic life event. Statistically significant ($p<0.05$) associations are in **bold**.

**Table S3.** Within-pair associations (Odds Ratio [OR] with 95% Confidence Intervals [CI]) of adulthood traumatic life events with depression (The Center for Epidemiological Studies-Depression [CES-D] ≥20) in depression discordant pairs by sex and zygosity in the Finnish Twin Cohort in 2011.

| Traumatic life events | ALL | DZ pairs | MZ pairs |
|---|---|---|---|
| | *N*<br>***OR***<br>*(95% CI)* | *N*<br>***OR***<br>*(95% CI)* | *N*<br>***OR***<br>*(95% CI)* |
| **Involved in a serious traffic accident** | | | |
| Male pairs | 132<br>1.23<br>(0.59, 2.56) | 85<br>1.13<br>(0.43, 2.92) | 47<br>1.40<br>(0.44, 4.41) |
| Female pairs | 245<br>1.00<br>(0.52, 1.92) | 167<br>1.30<br>(0.57, 2.96) | 78<br>0.63<br>(0.20, 1.91) |
| Homogeneity test for sex difference (p-value)* | 0.639 | 0.852 | 0.350 |
| **Involved in other serious accident** | | | |
| Male pairs | 127<br>2.00<br>(0.97, 4.12) | 84<br>**3.40**<br>**(1.25, 9.22)** | 43<br>0.83<br>(0.25, 2.73) |
| Female pairs | 240<br>1.21<br>(0.60, 2.46) | 162<br>1.27<br>(0.58, 2.80) | 78<br>1.00<br>(0.20, 4.95) |
| Homogeneity test for sex difference (p-value)* | 0.225 | 0.060 | 0.873 |
| **Witness to a fire or natural disaster** | | | |
| Male pairs | 130<br>1.00<br>(0.29, 3.45) | 87<br>1.00<br>(0.25, 4.00) | 43<br>1.00<br>(0.06, 16.0) |
| Female pairs | 243<br>1.17<br>(0.54, 2.52) | 165<br>0.80<br>(0.32, 2.03) | 78<br>3.00<br>(0.61, 14.9) |
| Homogeneity test for sex difference (p-value)* | 0.882 | 0.858 | 0.582 |
| **Injured by physical assault** | | | |
| Male pairs | 134<br>**2.33**<br>**(1.07, 5.09)** | 87<br>2.17<br>(0.82, 5.70) | 47<br>2.67<br>(0.71, 10.1) |

Table continues

| Traumatic life events | ALL | DZ pairs | MZ pairs |
|---|---|---|---|
| | *N*<br>***OR***<br>*(95% CI)* | *N*<br>***OR***<br>*(95% CI)* | *N*<br>***OR***<br>*(95% CI)* |
| Female pairs | 246<br>**3.33**<br>**(1.75, 6.35)** | 165<br>**3.43**<br>**(1.48, 7.96)** | 81<br>**3.20**<br>**(1.17, 8.73)** |
| Homogeneity test for sex difference (p-value)* | 0.531 | 0.537 | 0.846 |
| **Victim of sexual assault** | | | |
| Male pairs | 133<br>2.50<br>(0.49, 12.9) | 88<br>4.00<br>(0.45, 35.8) | 45<br>1.00<br>(0.06, 16.0) |
| Female pairs | 245<br>**2.94**<br>**(1.67, 5.18)** | 163<br>**2.82**<br>**(1.42, 5.61)** | 82<br>**3.2**<br>**(1.17, 8.74)** |
| Homogeneity test for sex difference (p-value)* | 1.000 | 0.739 | 0.607 |
| **Victim of or witness to a violent crime** | | | |
| Male pairs | 135<br>3.00<br>(0.61, 14.9) | 89<br>3.00<br>(0.31, 28.8) | 46<br>3.00<br>(0.31, 28.8) |
| Female pairs | 248<br>**3.00**<br>**(1.19, 7.56)** | 166<br>**2.60**<br>**(0.93, 7.29)** | 82<br>5.00<br>(0.58, 42.8) |
| Homogeneity test for sex difference (p-value)* | 0.496 | 0.587 | 0.460 |

CES-D=The Center for Epidemiological Studies-Depression; OR=Odds Ratio measured with conditional logistic regression analysis; CI= Confidence Intervals; N=number of pairs; n/a= not applicable (Not able to be calculated.) *Matched-pair cohort method (csmatch test in Stata) for testing homogeneity of RR estimates between Male and Female pairs. Statistically significant ($p<0.05$) associations are in **bold**.

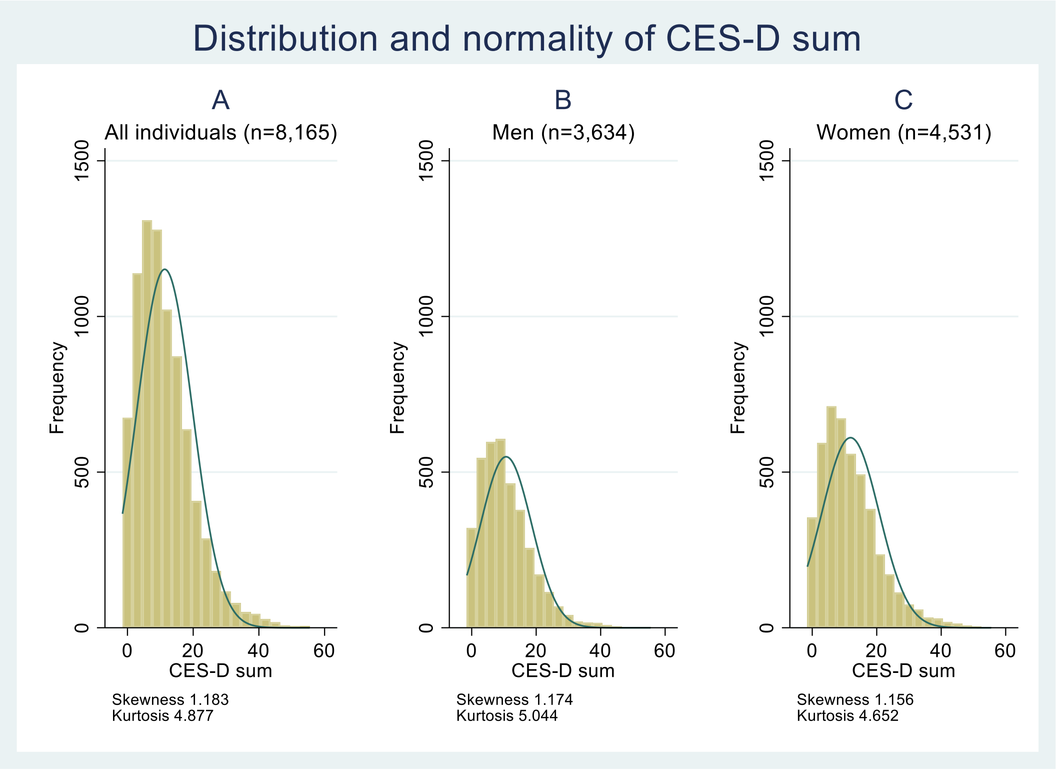


**Supplement Figure 1**. Distribution of the Center for Epidemiologic Studies Depression (CES-D) scale sum in the whole cohort (A) and separately in men (B) and in women (C).
